# Comparative Assessment of Myocardial Work Performance during Spontaneous Rhythm, His Bundle Pacing, and Left Bundle Branch Area Pacing: Insights from the EMPATHY Study

**DOI:** 10.1101/2023.07.14.23292694

**Authors:** Giorgia Azzolini, Nicola Bianchi, Francesco Vitali, Michele Malagù, Cristina Balla, Martina De Raffele, Matteo Bertini

## Abstract

**Background:** Physiological pacing has gained significant interest due to its potential to achieve optimal hemodynamic response. This study aimed to assess left ventricular performance in terms of electrical parameters, specifically QRS duration, and mechanical performance, evaluated as myocardial work. We compared His Bundle Pacing (HBP) and Left Bundle Branch Area Pacing (LBBAP) to evaluate their effects.

**Methods:** Twenty-four patients with class I or IIa indications for pacing were enrolled in the study, with 12 patients undergoing HBP implantation and another 12 patients undergoing LBBAP implantation. A comprehensive analysis of myocardial work was conducted.

**Results:** Our findings indicate that there were no major differences in terms of spontaneous and HBP activation in myocardial work, except for global wasted work (217 mmHg% vs. 283 mmHg%; p 0.016) and global work efficiency (87 mmHg% vs. 82 mmHg%; p 0.049). There were no significant differences observed in myocardial work between spontaneous activation and LBBAP. Similarly, no significant differences in myocardial work were found between HBP and LBBAP.

**Conclusions:** Both pacing modalities provide physiological ventricular activation without significant differences when compared to each other. Moreover, there were no significant differences in QRS duration between HBP and LBBAP.

However, LBBAP demonstrated advantages in terms of feasibility, as it achieved better lead electrical parameters compared to HBP (threshold@0.4 ms 0.6 V vs. 1 V; p=0.045. Sensing 9.4 mV vs. 2.4 mV; p<0.001). Additionally, LBBAP required less fluoroscopy time (6 min vs. 13 min; p=0.010) and procedural time (81 min vs. 125 min; p=0.004) compared to HBP.

**Clinical Perspective:** *What is Known:* His Bundle Pacing (HBP) and Left Bundle Branch Area Pacing (LBBAP) have been recognized as more physiological alternatives to traditional right ventricular pacing. LBBAP has shown greater feasibility compared to HBP, although direct comparison data between the myocardial work in HBP and LBBAP are limited.

*What the Study Adds:* Our study contributes to the existing knowledge by demonstrating that both HBP and LBBAP provide physiological ventricular activation, with no significant differences observed between the two pacing modalities in terms of myocardial work and QRS duration. However, LBBAP showcased advantages such as reduced need for fluoroscopy, shorter procedural time, and improved electrical parameters. These findings further support the potential of LBBAP as a favorable pacing option. Graphical abstract

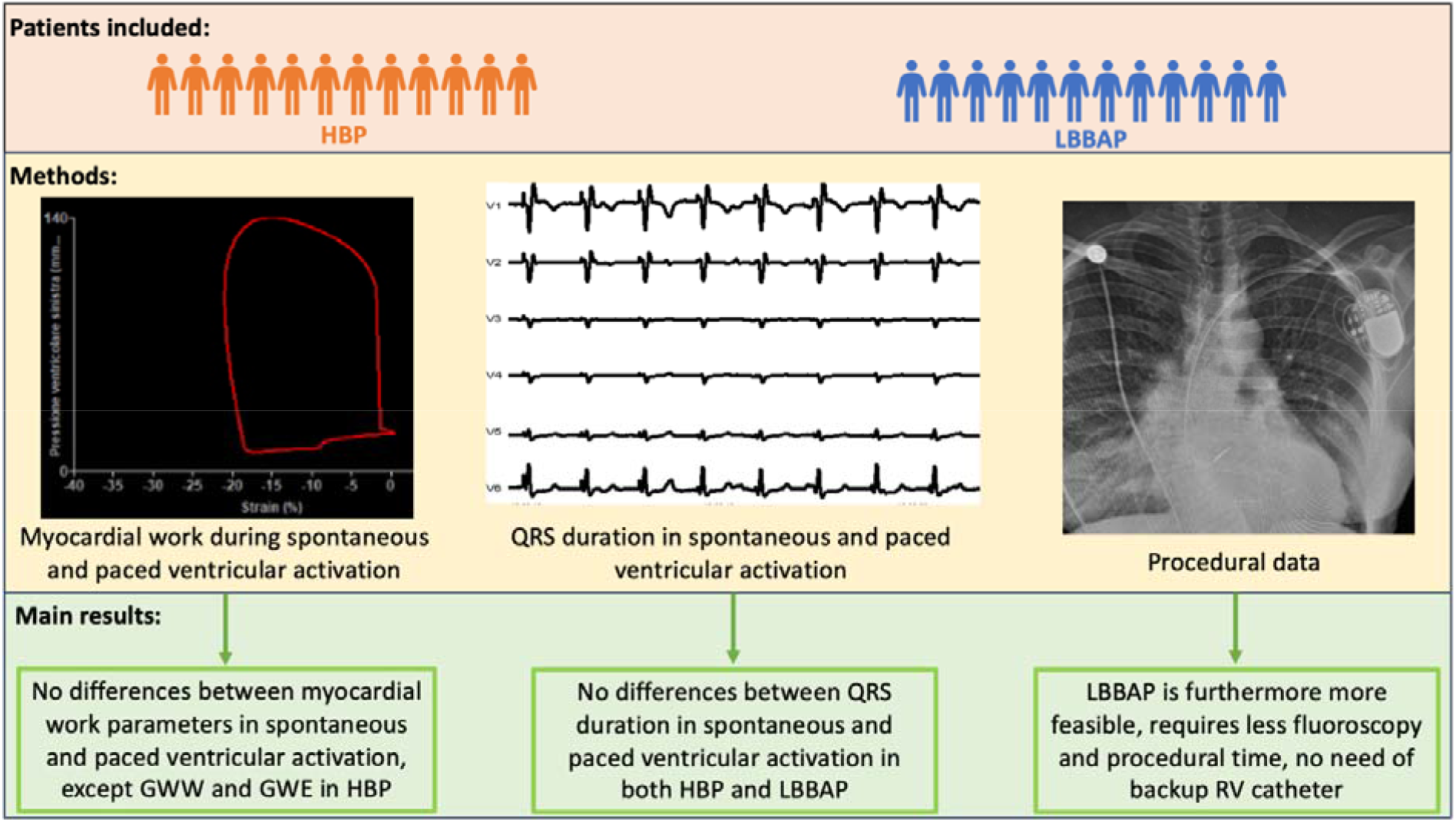

## Background

Historically, dual-chamber pacing emerged as a superior alternative to single-chamber pacing due to its ability to maintain physiological atrioventricular synchrony whenever possible. However, the detrimental effects of mid– and long-term right ventricular (RV) pacing [1–4] have underscored the necessity for an alternative pacing site to achieve more physiological ventricular activation. Cardiac resynchronization therapy (CRT) using biventricular pacing partially addressed this issue [5]. Nonetheless, biventricular pacing still results in non-physiological activation, as it originates from the epicardium and propagates through the myocardium instead of the conduction system. In recent years, conduction system pacing (CSP) has garnered substantial attention and adoption in clinical practice [6,7]. Initially, the main focus of CSP was His Bundle Pacing (HBP) [8,9]. However, Left Bundle Branch Area Pacing (LBBAP) has emerged as a promising alternative, surpassing many of the limitations associated with HBP in terms of feasibility, electrical parameters, and device settings [10–12]. Although no direct comparison data between myocardial work in HBP and LBBAP are available. In this study, we aimed to investigate the effects of HBP and LBBAP on left ventricular (LV) performance in a subset of patients with class I or IIa indications for pacemaker (PM) implantation.

## Methods

This study is derived from the EMPATHY study [13], which is a prospective, single-center cohort study conducted at the Cardiology Unit of Azienda Ospedaliero-Universitaria di Ferrara, Italy. The study enrolled consecutive patients who underwent HBP or LBBAP. The study protocol was registered on ClinicalTrials.gov (NCT05222672) and received approval from the local ethics committee. Informed consent was obtained from all patients.

The inclusion criteria for the study were as follows: (1) class I or IIa indication for pacemaker implantation, based on the European guidelines [14]; (2) age ≥18 years; (3) signed written informed consent. The exclusion criteria included: (1) inability to provide informed consent; (2) pregnancy; (3) severe mitral or aortic valve disease; (4) left ventricular ejection fraction (LVEF) <35%.

The primary endpoint of the study was to compare LV myocardial work during spontaneous ventricular activation (SVA) with HBP or LBBAP. The secondary endpoint was to compare the change in LV myocardial work between stimulated activation with HBP and LBBAP, as well as SVA.

### Implantation

#### His bundle pacing

The index procedure involved the implantation of a pacemaker and simultaneous three-dimensional electroanatomical Mapping (3D-EAM). The right ventricle and the His bundle area were mapped non-fluoroscopically using a high-density mapping catheter, which was inserted via the femoral vein. The pacing leads were inserted via the left cephalic or axillary vein and positioned using a combination of 3D-EAM and fluoroscopy [13].

For His bundle pacing an active fixation lead (SelectSecure 3830, Medtronic, Minneapolis, MN, USA, or Solia S, Biotronik, Berlin, Germany) was placed using a non-deflectable sheath (C315, Medtronic, Minneapolis, MN, USA, or Selectra 3D, Biotronik, Berlin, Germany). The position of the His bundle lead was confirmed using unipolar and bipolar intracardiac electrograms, with standard criteria employed to determine selective and non-selective His capture [15]. The pacing threshold was defined as the intraprocedural unipolar capture threshold at 0.4 ms during the implant procedure. In all patients, a backup right ventricular lead was implanted, and an atrial lead was added, if necessary, based on the pacing indication (figure 1). Antibiotic prophylaxis and antithrombotic drugs were administered following the protocols of the center and international guidelines [16,17].

**Figure 1:**
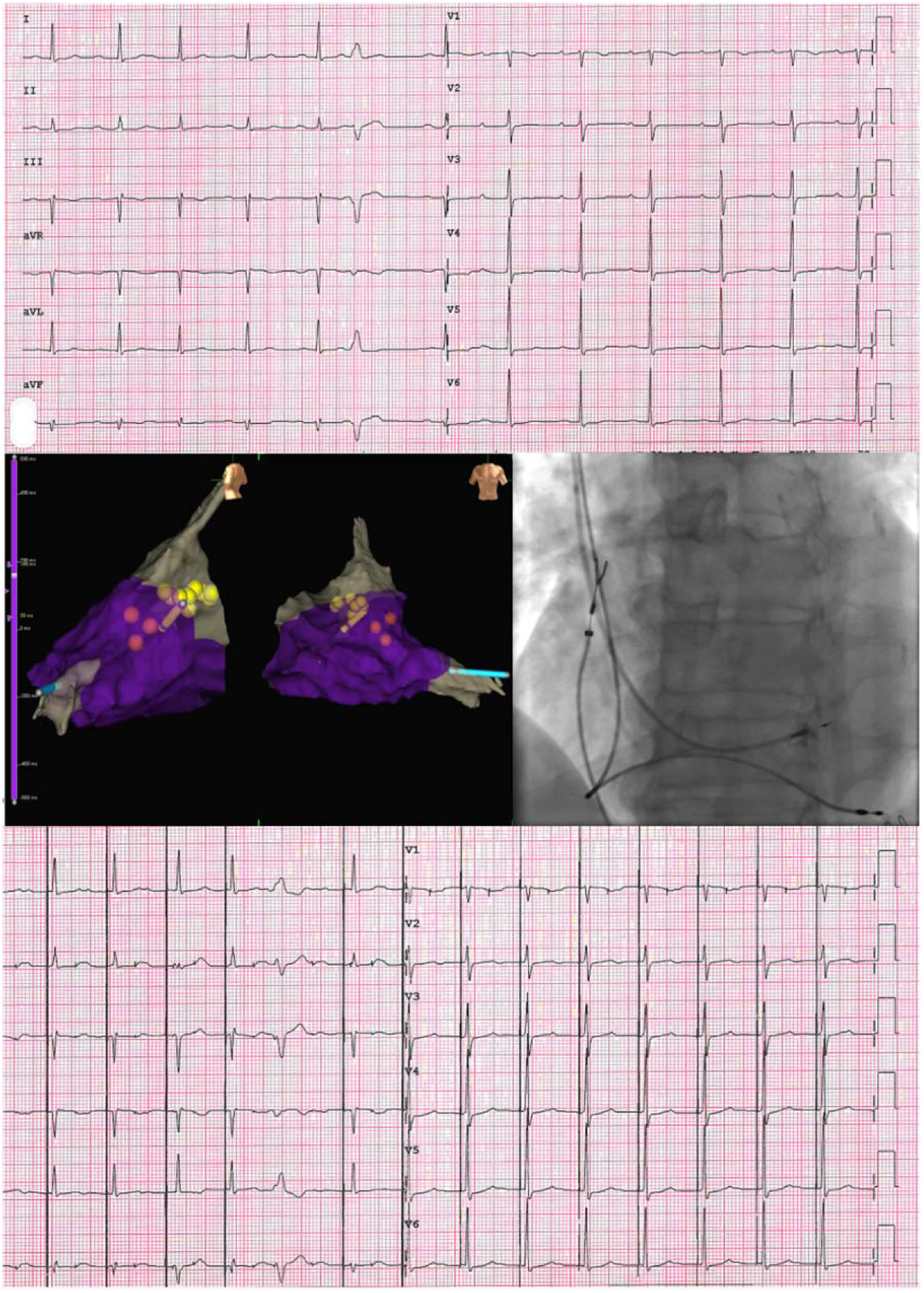
The upper part of the image shows the spontaneous ventricular activation as a twelve-lead ECG. In the central part, on the left, a 3DEAM of the RV is displayed, with highlighted yellow dots representing the His bundle. The yellow catheter corresponds to the His bundle lead, while the light blue catheter represents the RV back-up lead. In the central part, on the right, a fluoroscopic anterior-posterior view of the His lead and the RV back-up lead is shown. In the inferior part of the image, an ECG with selective HBP is show.

### Left bundle branch area pacing

Before the procedure, the thickness of the basal interventricular septum and the presence of septal scar were assessed. The pacing leads were inserted via the left cephalic or axillary vein. The active fixation 3830 lumenless lead (Medtronic, Minneapolis, USA) and the non-deflectable C315His sheath, as well as the Biotronik Solia 60 stylet-driven lead (Biotronik, Berlin, Germany) and Selectra 3D, were used as the pacing leads and delivery guiding catheters.

The procedure began by advancing the guiding catheter into the right ventricle (RV) over a J wire. In the left anterior oblique (LAO) view (30-40°), contrast injection via the guiding catheter was performed to delineate the septum. After counterclockwise rotation of the guiding catheter to achieve a perpendicular orientation to the septum, a pace map was conducted by exposing the tip of the pacing lead [18]. The aim was to obtain a QRS morphology in lead II that was more positive than in lead III, aVr and aVl discordant, and W’ pattern with a notch at the nadir of QRS complex in V1.

The screwing technique varied depending on the type of lead used. For lumenless leads, 3-5 rapid rotations were applied based on the thickness of the ventricular septum while keeping the guiding catheter in position. For stylet-driven leads, the first screw was exposed, and after a pacing check, 2-3 rapid rotations similar to lumenless leads were performed. As the screw advanced into the interventricular septum, the paced QRS morphology exhibited a right bundle branch block (RBBB) pattern, with the notch at the nadir of the QRS in lead V1 shifting to the end of the QRS, indicating successful LBBAP (figure 2). A fast LV activation time in V6 <75 ms was also aimed for [19]. The myocardial current of injury (COI) was recorded in the electrogram (EGM). In cases where perforation into the LV occurred, the lead was repositioned at a different location. An atrial lead was implanted when needed based on the pacing indication. Antibiotic prophylaxis and antithrombotic drugs were administered following the center’s protocols [16,17].

**Figure 2:**
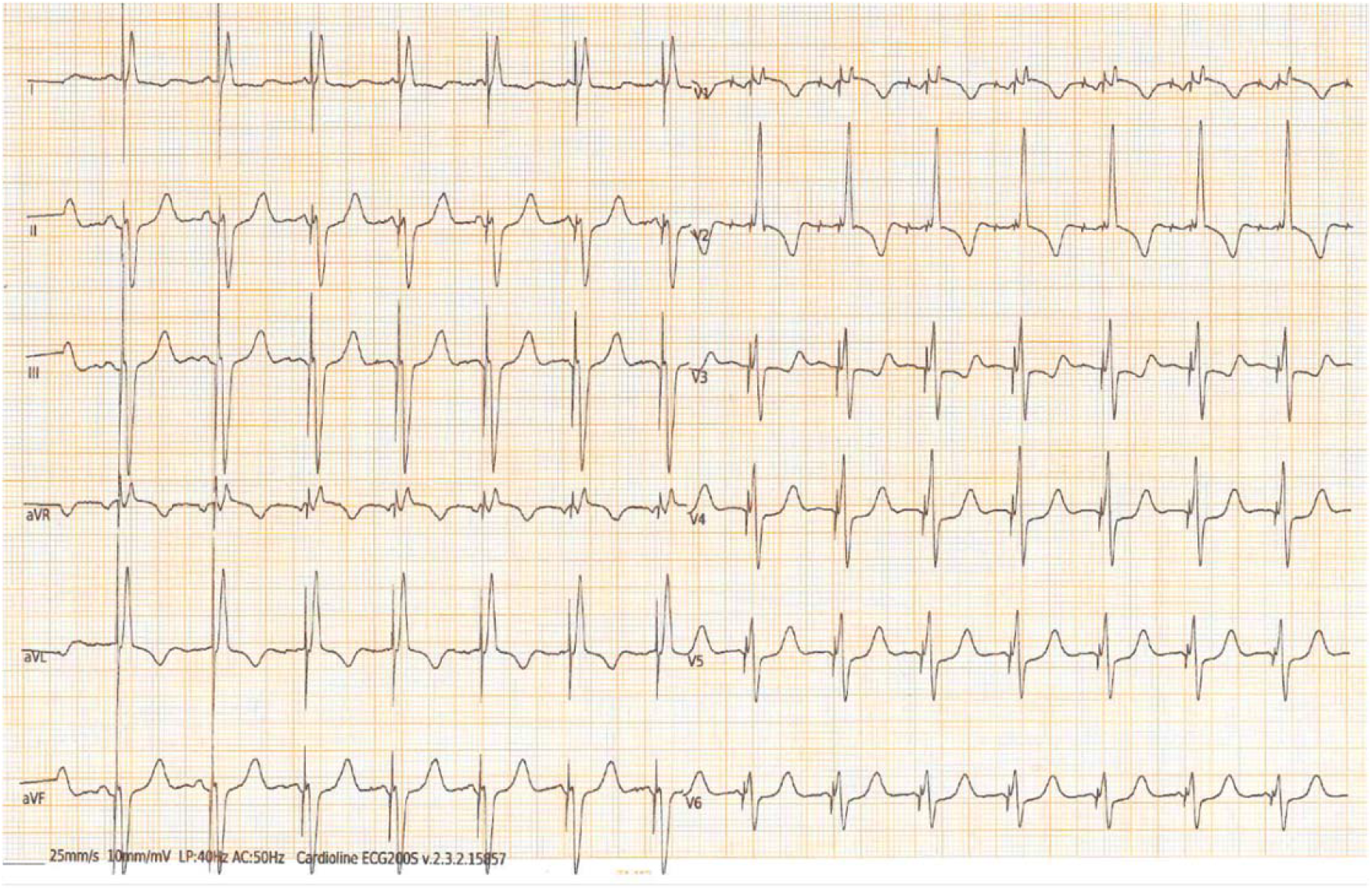
The twelve-lead ECG recorded during LBBAP (unipolar output 1 V / 0.4 ms) demonstrates a paced QRS complex with a right bundle branch morphology pattern, characterized by a terminal R/r’ wave in lead V1. The QRS complex duration is narrow, measuring 105 ms, and the LVAT (left ventricular activation time) is 67 ms.

During the procedure, the pacing threshold was measured in the unipolar configuration at 0.4 ms, and the impedance was determined in the unipolar configuration as well. The final R-wave amplitude was measured in the bipolar configuration. Successful LBBAP was considered when a deep septal lead position was achieved and the paced QRS complex included a terminal R/r wave in lead V1, indicating a delay in right ventricle activation. In rare cases, a QS configuration (lack of terminal R) in V1 was accepted if a terminal R/r wave in lead V1 appeared during programmed stimulation or other features indicating LBBAP, as described below, were present.

### Imaging and electrocardiogram

After the procedure, all patients underwent echocardiographic evaluation to assess various parameters. Basic information such as LV volume (in mL), LV ejection fraction (LVEF) (in %), atrial volume (in mL), and the degree of valve diseases were collected. Global longitudinal strain (GLS) and myocardial work (MW) were analyzed as part of the evaluation. MW is a novel echocardiographic technique that utilizes speckle tracking analysis to estimate left ventricular performance by measuring the area under the pressure-strain loops curve derived from GLS and blood pressure [20–22]. All echocardiographic examinations were performed using the GE Vivid E9 with M5S transducers, and GLS and MW analyses were conducted offline using EchoPAC software V.202 (GE Healthcare, Chicago, IL, USA). The echocardiographic parameters were assessed according to international standards [23]. The collected MW parameters included:

(i) Global constructive work (GCW): The sum of work performed during myocardial shortening in systole and myocardial lengthening during isovolumetric diastole.
(ii) Global wasted work (GWW): The sum of work performed by myocardial lengthening in systole and myocardial shortening during isovolumetric diastole.
(iii) Global work index (GWI): The work performed throughout systole, specifically between mitral valve closure and opening.
(iv) Global work efficiency (GWE): Expressed as the percentage ratio of GCW to the sum of GCW and GWW.

A twelve-lead ECG was performed and analyzed as well. Echocardiography and ECG evaluations were conducted in different conditions, including SVA and during HBP or LBBAP, respectively. The measurements for HBP and LBBAP were obtained during pacing at a fixed rate in DDD mode with an optimized AV delay or in VVI mode, depending on whether the patients were in sinus rhythm or atrial fibrillation

### Statistical analysis

Continuous variables were expressed as mean ± standard deviation when normally distributed, as estimated using the Shapiro–Wilk test, or as median and interquartile range. All variables were not normally distributed. Categorical variables were expressed as number and percentages.

Differences between repeated measurements during SVA and HBP or LBBAP were assessed using the Wilcoxon rank-sum test. Differences between independent measurement in HBP and LBBAP were assessed using the Mann-Whitney U test. In order to adjust for baseline parameter during SVA the comparison between myocardial work during HBP and LBBAP was not done between absolute values, but between relative values calculated as difference between stimulated and spontaneous parameter divided by the spontaneous one [example: dGCW_LBBAP_ = (GCW_LBBAP_-GCW_SVA_)/GCW_SVA_] and multiplied for 100 to obtain percentage change.

*p* values < 0.05 were considered statistically significant. The statistical analysis was performed using STATA, version 15.0 (StataCorp LLC, Texas US).

## Results

A total of twenty-four patients were enrolled in the study, with four of them being female (two in each group). The patients were matched for comorbidities and implant characteristics. The baseline clinical characteristics, procedural data, and electrical parameters are summarized in Table 1. The median age was 79 years (IQR 73-85) in the HBP group and 81 years (IQR 73-85) in the LBBAP group, with no significant difference between the groups. There were no significant differences between the groups in terms of the main cardiovascular risk factors such as arterial hypertension, dyslipidemia, and diabetes (Table 1).

**Table 1.** Baseline characteristics of the patients included in the study. Procedural data and electrical parameters after implantation. Data expressed as number (%) or median (interquartile range). BMI: body mass index; CAD: coronary artery disease; EF: ejection fraction.

|  | ***HBP***<br>***(12 patients)*** | ***LBBAP***<br>***(12 patients)*** | ***p-value*** |
| --- | --- | --- | --- |
| **Baseline characteristics** |  |  |  |
| Age (years) | 79 (73-85) | 81 (73-85) | 0.88 |
| Female sex | 2 (17) | 2 (17) | 1.00 |
| BMI (kg/m<sup>2</sup>) | 25.1 (23.4-28.3) | 24.7 (22.3-29.1) | 0.69 |
| CAD | 4 (33) | 2 (17) | 0.64 |
| Atrial Fibrillation | 7 (58) | 5 (42) | 0.68 |
| Diabetes mellitus | 0 (0) | 4 (33) | 0.093 |
| Hypertension | 10 (83) | 9 (75) | 1.00 |
| Dyslipidemia | 6 (50) | 8 (67) | 0.68 |
| **Procedural data** |  |  |  |
| Procedure duration (min) | 125 (120-140) | 81 (70-120) | 0.004* |
| Fluoroscopy time (min) | 13 (9-21) | 6 (5-11) | 0.010* |
| Selective HPB capture | 6 (50) | - | - |
| **Electrical parameters** |  |  |  |
| Lead impedance (ohm) | 509 (447-568) | 718 (609-795) | 0.002* |
| Lead sensing (mV) | 2.4 (1.8-4.8) | 9.4 (8-13.5) | <0.001* |
| Lead threshold (V@0,4 ms) | 1.0 (0.5-2.5) | 0.6 (0.4-0.8) | 0.045* |
| EF (%) | 51 (42-63) | 55 (51-56) | 0.750 |

Furthermore, no significant differences were observed in the major cardiovascular comorbidities that could affect the implantation procedure. Specifically, there were no significant differences in the history of atrial fibrillation (AF) (58% in the HBP group vs. 42% in the LBBAP group; p=0.68) or coronary artery disease (CAD) (33% in the HBP group vs. 17% in the LBBAP group; p=0.64). Baseline LVEF were also comparable between the two groups (51% in the HBP group vs. 55% in the LBBAP group; p=0.75).

Selective HBP was obtained in six patients (50%). LBBAP was feasible in all the patients. The procedural duration was significantly longer in the HBP group compared to the LBBAP group (125 minutes; IQR 120-140 vs. 81 minutes; IQR 70-120; p 0.004). Similarly, intraprocedural fluoroscopy time was higher in the HBP group compared to the LBBAP group (13 minutes; IQR 9-21 vs. 6 minutes; IQR 5-11; p 0.01). Electrical lead parameters were assessed after implantation. Lead impedance was significantly lower in the HBP group compared to the LBBAP group (509 ohm; IQR 447-568 vs. 718 ohm; IQR 608-795; p 0.002). Lead bipolar sensing was significantly lower in the HBP group compared to the LBBAP group (2.4 mV; IQR 1.8-4.8 vs. 9.5 mV; IQR 8-13.5; p < 0.001).

Lead unipolar threshold was significantly higher in the HBP group compared to the LBBAP group (1.0 V; IQR 0.5-2.5 vs. 0.6 V; IQR 0.4-0.8; p 0.045).

### ECG and SVA myocardial work

The baseline data for electrocardiographic and myocardial work are presented in Table 2. The spontaneous QRS duration did not show a statistically significant difference between the HBP and LBBAP groups (106 ms; IQR 88-140 vs. 115 ms; IQR 90-132, p=0.76). Similarly, the paced QRS duration did not exhibit a statistically significant difference between the HBP and LBBAP groups (124 ms; IQR 98-140 vs. 128 ms; IQR 118-136, p=0.62).

**Table 2.** Baseline electrocardiographic and myocardial work parameters in spontaneous ventricular activation. Data expressed as median (interquartile range). GWI: global work index; GCW: global constructive work; GWW: global wasted work; GWE global work efficiency.

|  | ***HBP***<br>***(12 patients)*** | ***LBBAP***<br>***(12 patients)*** | ***p-value*** |
| --- | --- | --- | --- |
| Spontaneous QRS duration (ms) | 106 (88-140) | 115 (90-132) | 0.76 |
| Paced QRS duration (ms) | 124 (98-140) | 128 (118-136) | 0.62 |
| Spontaneous GWI (mmHg%) | 1120 (902-1763) | 1548 (1072-2273) | 0.094 |
| Spontaneous GCW (mmHg%) | 1648 (1044-2152) | 2089 (1705-2594) | 0.052 |
| Spontaneous GWW (mmHg%) | 217 (125-249) | 137 (105-286) | 0.670 |
| Spontaneous GWE (mmHg%) | 87 (80-90) | 93 (83-95) | 0.065 |

There were no significant differences observed in any of the myocardial work indexes between the HBP and LBBAP groups.

In the first part of the analysis, a comparison was made between SVA and paced ventricular activation in the HBP group (Table 3). No significant differences were found in the GWI (1110 mmHg% IQR 902-1763 in SVA vs. 1020 mmHg% IQR 822-1969 in HBP; p 0.534) and GCW (1648 mmHg% IQR 1044-2152 in SVA vs. 1505 mmHg% IQR 1151-2133 in HBP; p 0.075) between the two types of activation.

**Table 3.** Difference between myocardial work parameters in spontaneous ventricular and paced ventricular activation in HBP and LBBAP. Data expressed as median (interquartile range). GWI: global work index; GCW: global constructive work; GWW: global wasted work; GWE global work efficiency.

| **HBP (12 patients)** |  |  |  |
| --- | --- | --- | --- |
|  | ***Spontaneous*** | ***Paced*** | ***p-value*** |
| QRS duration (ms) | 106 (88-140) | 124 (98-140) | 0.75 |
| GWI (mmHg%) | 1110 (902–1763) | 1020 (822-1969) | 0.534 |
| GCW (mmHg%) | 1648 (1044-2152) | 1505 (1151-2133) | 0.075 |
| GWW (mmHg%) | 217 (125-249) | 283 (205-354) | 0.016* |
| GWE (mmHg%) | 87 (80-90) | 82 (73-90) | 0.049* |
| **LBBAP (12 patients)** |  |  |  |
|  | ***Spontaneous*** | ***Paced*** | ***p-value*** |
| QRS duration (ms) | 115 (90-132) | 128 (118-136) | 0.05 |
| GWI (mmHg%) | 1548 (1072-2273) | 1545 (1363-1961) | 0.374 |
| GCW (mmHg%) | 2089 (1705-2594) | 2320 (2073-2530) | 0.929 |
| GWW (mmHg%) | 137 (105-286) | 264 (195-341) | 0.091 |
| GWE (mmHg%) | 93 (83-95) | 87 (83-92) | 0.109 |

However, GWW was significantly higher in HBP ventricular activation compared to SVA (217 mmHg% IQR 125-249 in SVA vs. 283 mmHg% IQR 205-354 in HBP; p-value = 0.016), and GWE was significantly lower in HBP ventricular activation compared to SVA (87% IQR 80-90 in SVA vs. 82% IQR 73-90 in paced rhythm; p 0.049).

In the second part of the analysis, a comparison was made between SVA and paced ventricular activation in the LBBAP group (Table 3). No differences were found in myocardial work parameters between SVA and paced rhythm. In particular, GWI was 1548 (1072-2273) mmHg% in SVA vs 1545 (1363-1961) mmHg% in paced rhythm with p-value=0,374; GCW was 2089 (1705-2594) mmHg% in SVA vs 2320 (2073-2530) mmHg% in paced rhythm with p-value=0,929; GWW was 137 (105-286) mmHg% in SVA vs 264 (195-341) mmHg% in paced rhythm with p-value=0,091; GWE was 93 (83-95)% in SVA vs 87 (83-92)% in paced rhythm with p-value=0,109 (Figure 2).

Relative differences between SVA and paced ventricular activation of all myocardial work indexes and paced QRS duration did not show significant differences between the LBBAP and HBP groups. The relative differences in GWI, GCW, GWW, GWE and paced QRS duration were comparable between the two groups (Table 4).

**Table 4.** Comparison of HBP and LBBAP QRS duration and parameters of myocardial work as relative differences between paced and spontaneous ventricular activation. Data expressed as median (interquartile range). dQRS: relative difference in QRS [(QRS_paced_– QRS_spontaneus_)*100/QRS_spontaneus_]; dGWI: relative difference in global work index [(GWI_paced_– GWI_spontaneus_)*100/GWI_spontaneus_]; GCW: global constructive work [(GCW_paced_– GCW_spontaneus_)*100/GCW_spontaneus_]; GWW: global wasted work [(GWW_paced_– GWW_spontaneus_)*100/GWW_spontaneus_]; GWE global work efficiency [(GWE_paced_– GWE_spontaneus_)*100/GWE_spontaneus_].

|  | ***HBP***<br>**(12 patients)** | ***LBBAP***<br>**(12 patients)** | ***p-value*** |
| --- | --- | --- | --- |
| dQRS (%) | 0 (-7.5 - +42.8) | +16.4 (-4.9 - +32.0) | 0.54 |
| dGWI (%) | +0.1 (-18.4 - +36.4) | -6.3 (-22.6 - +24.4) | 0.67 |
| dGCW (%) | 12.3 (-2.8 - +16.2) | -0.9 (-10.2 - +6.2) | 0.25 |
| dGWW (%) | +66.4 (+8.4 - +91.7) | +113.3 (-5.3 - +218.2) | 0.53 |
| dGWE (%) | -2.4 (-16.7 - +1.1) | -5.3 (-9.7 - +1.1) | 0.77 |

## Discussion

Conduction system pacing, which includes HBP and LBBAP is a physiological pacing modality that stimulates the myocardium through specialized conduction fascicles [6]. It aims to avoid the electrical and mechanical dyssynchrony caused by the traditional RV Apical Pacing and its associated detrimental effects.

In our study, we assessed the impact of HBP and LBBAP on left ventricular performance, focusing on electrical and mechanical synchronization, in a subgroup of patients with a class I or IIa indication for pacemaker implantation. Our main findings are as follows:

(i) There were no statistically significant differences in terms of myocardial work index and global constructive work between spontaneous and paced activation in both LBBAP and HBP groups.
(ii) The relative difference in all myocardial work parameters between sinus and paced activation did not show statistically significant differences between the two groups. Spontaneous and paced QRS durations were not statistically different in either group.
(iii) The relative difference between spontaneous and paced QRS durations did not exhibit statistically significant differences between the HBP and LBBAP groups.

Myocardial work (MW) is an emerging tool in studying myocardial mechanics [20]. Unlike traditional parameters such as ejection fraction, MW incorporates both deformation and load, providing additional information on cardiac performance. It is also more effective in quantitatively assessing mechanical synchrony and efficiency compared to speckle tracking imaging [21,22]. Furthermore, MW offers a better evaluation of synchronization than electrocardiographic features, such as QRS duration [24]. In our study, we found no significant differences in the global work index and global constructive work between spontaneous and paced ventricular activation in both groups. HBP exhibits a significantly higher global wasted work and lower global work efficiency compared to SVA, likely attributed to the considerable percentage of non-selective HBP. In our study group, selective HBP was achieved in half of the patients, aligning with previous literature. [25]. Furthermore, the comparison of relative differences between spontaneous and paced ventricular activation in all myocardial work parameters did not reveal any statistically significant differences between the two groups. Additionally, in the LBBAP group, global wasted work and global work efficiency were comparable between spontaneous and paced rhythm. The Empathy study [13] demonstrated that HBP had similar myocardial performance to SVA and superior ventricular efficiency compared to RV Apical Pacing. A recent study by Wang et al. [26] showed that LBBAP resulted in more effective myocardial work than RVP.

The duration of surface QRS has commonly been used as an approximate measure of electrical synchronization [27]. In our study, we found no significant differences in QRS duration between spontaneous and paced ventricular activation in both the HBP and LBBAP groups, as well as between paced activation in the two groups. These results align with previous studies [8, 9, 13, 28] that have reported similar findings regarding QRS duration in relation to different pacing modalities.

HBP is a technically challenging procedure that requires time and the use of a backup right ventricular pacing lead [18]. The success rate of HBP implantation varies in the literature [25, 29–30], but it generally improves with increasing experience in both HBP and LBBAP [28]. While early studies reported longer fluoroscopy and procedural times for HBP, recent experiences have shown reduced times, although still longer than traditional RVP procedures [25]. In our study population, both procedural and fluoroscopy times were longer in the HBP group compared to the LBBAP group (125 vs 80, p=0.004; 13 vs 6, p=0.01, respectively), despite the use of 3D-EAM during HBP.

Another important aspect to consider is the type of capture achieved during HBP. It can be either selective, capturing only the His bundle tissue, or non-selective, involving fusion capture of the His bundle and adjacent myocardium [8]. The type of capture depends on the pacing location, surrounding atrial or ventricular tissue, and pacing output amplitude [25]. Achieving selective HBP is not always straightforward in our study population, as it was achieved in only six patients (50%) in line with previous literature [25].

In contrast, LBBAP is defined as capturing the subendocardial area on the left side of the interventricular septum, with or without simultaneous conduction system capture. It encompasses techniques such as Left Bundle Branch Pacing (LBBP), Left Fascicular Pacing (LFP), and Left Ventricular Septal Pacing (LVSP) [18]. Consequently, LBBAP is more predictable and easier to achieve compared to HBP, with a higher implant success rate [10, 12, 19]. Another advantage of LBBAP is that the lead is positioned deep in the ventricular septal myocardium, providing backup septal pacing in case of loss of left bundle branch capture due to more distal conduction system disease.

The electrical performance of the implanted ventricular leads differed significantly between the two groups, with the HBP group exhibiting higher impedance, lower sensing, and higher unipolar threshold compared to the LBBAP group. These findings emphasize the clinical advantages of LBBAP over HBP, as LBBAP consistently exhibits superior electrical parameters

### Limitations

One of the limitations of our study is the relatively small sample size, with only 24 patients. However, given the scarcity of data comparing HBP and LBBAP in terms of myocardial performance, our findings provide valuable insights. Another limitation is that our study was conducted at a single center, which may introduce a potential limitation in terms of generalizability. However, this limitation also eliminates the potential bias associated with variations in expertise between different centers. Additionally, the assessment of mechanical synchrony was dependent on the quality of echocardiography images. Nevertheless, the use of speckle tracking technology has been shown to reduce this limitation, as it has a feasibility rate of over 90% [31].

## Conclusions

Both His Bundle Pacing and Left Bundle Branch Area Pacing offer physiological ventricular activation with optimal electrical and mechanical synchrony. However, LBBAP emerges as a more favorable option due to its greater feasibility, reduced need for fluoroscopy, and shorter procedural time, effectively addressing the limitations associated with HBP.

## Data Availability

All the data are available

